# Implementation and Assessment of the HIV Enhanced Access Testing in the Emergency Department (HEATED) Program in Nairobi, Kenya: A Quasi-Experimental Prospective Study

**DOI:** 10.1101/2024.04.03.24305277

**Authors:** Adam R. Aluisio, Joshua Smith-Sreen, Agatha Offorjebe, Wamutitu Maina, Sankei Pirirei, John Kinuthia, David Bukusi, Harriet Waweru, Rose Bosire, Daniel K. Ojuka, McKenna C. Eastment, David A. Katz, Michael J. Mello, Carey Farquhar

**Author notes:** **Corresponding author:** Adam R. Aluisio, MD, MSc, DTM&H, Department of Emergency Medicine, Alpert Medical School of Brown University, 55 Claverick Street, Room 274, Providence, RI 02903.

## Abstract

**Background:** Persons seeking emergency injury care are often from underserved key populations (KPs) and priority populations (PPs) for HIV programming. While facility-based HIV Testing Services (HTS) in Kenya are effective, emergency department (ED) delivery is limited, despite the potential to reach underserved persons.

**Methods:** This quasi-experimental prospective study evaluated implementation of the HIV Enhanced Access Testing in Emergency Departments (HEATED) at Kenyatta National Hospital ED in Nairobi, Kenya. The HEATED program was designed using setting specific data and utilizes resource reorganization, services integration and HIV sensitization to promote ED-HTS. KPs included sex workers, gay men, men who have sex with men, transgender persons and persons who inject drugs. PPs included young persons (18-24 years), victims of interpersonal violence, persons with hazardous alcohol use and those never previously HIV tested. Data were obtained from systems-level records, enrolled injured patient participants and healthcare providers. Systems and patient-level data were collected during a pre-implementation period (6 March - 16 April 2023) and post-implementation (period 1, 1 May - 26 June 2023). Additional, systems-level data were collected during a second post-implementation (period 2, 27 June – 20 August 2023). Evaluation analyses were completed across reach, effectiveness, adoption, implementation and maintenance framework domains.

**Results:** All 151 clinical staff were reached through trainings and sensitizations on the HEATED program. Systems-level ED-HTS increased from 16.7% pre-implementation to 23.0% post-implementation periods 1 and 2 (RR=1.31, 95% CI:1.21-1.43; p<0.001) with a 62.9% relative increase in HIV self-test kit provision. Among 605 patient participants, facilities-based HTS increased from 5.7% pre-implementation to 62.3% post-implementation period 1 (RR=11.2, 95%CI:6.9-18.1; p<0.001). There were 440 (72.7%) patient participants identified as KPs (5.6%) and/or PPs (65.3%). For enrolled KPs/PPs, HTS increased from 4.6% pre-implementation to 72.3% post-implementation period 1 (RR=13.8, 95%CI:5.5-28.7, p<0.001). Systems and participant level data demonstrated successful adoption and implementation of the HEATED program. Through 16-weeks post-implementation a significant increase in ED-HTS delivery was maintained as compared to pre-implementation.

**Conclusions:** The HEATED program increased ED-HTS and augmented delivery to KPs/PPs, suggesting that broader implementation could improve HIV services for underserved persons, already in contact with health systems.

## INTRODUCTION

There are approximately 38 million people living with HIV (PLH) globally, of whom 70% reside in sub-Saharan Africa.^1,2^ Although progress was been made toward the UNAIDS 95-95-95 HIV control targets, they are at risk of not being achieved.^3^ In sub-Saharan Africa, Kenya has reduced the national HIV prevalence, however incidence reduction targets have not been met, and in 2021 there was an 7.8% increase in new infections.^4^ The epidemic in Kenya is driven by higher-risk persons who have been insufficiently reached and underserved by HIV Testing Services (HTS).^4–6^ These include key populations (KPs) of sex workers (SWs), men who have sex with men (MSM), transgender persons and people who inject drugs (PWID) in which HIV prevalence is five-fold higher than the general population.^5,7^ Additional priority populations (PPs) including men, young people (<25 years) and persons who use drugs have been insufficiently reached further contributing to epidemic propagation.^8–11^ Young people in Kenya account for 42% of new infections and one in four Kenyan men with HIV are undiagnosed.^5^ To achieve epidemic control, these higher-risk persons must be reached for HIV testing and linkage to care.^2,9^ However with persistent HIV programming barriers stemming from structural and cultural aspects as well as stigma and discrimination,^12–16^ innovative service delivery approaches are needed.

Emergency Departments (ED) in low- and middle-income countries (LMICs) are an under-used HTS service delivery point with the potential to reach KP/PPs already in contact with health systems.^17,18^ In LMICs, EDs provide care to large numbers of patients who may not otherwise regularly access healthcare, and often for treatment of injuries.^17,19,20^ Data from sub-Saharan Africa shows patients seeking injury treatments have high HIV burdens and are often first diagnosed during those evaluations.^21–25^ Globally, young people and men are most likely to experience injuries,^26,27^ and KPs have elevated risks for interpersonal violence, gender-based violence, and self-harm.^28,29^ In Kenya, data from the National AIDS and STIs Control Programme (NASCOP) demonstrates high prevalence of violence experienced by SW (48%), PWID (44%), and MSM (20%).^30^ Given injury risk burdens among KP/PPs in Kenya and subsequent needs for emergency care, development of effective ED-HTS with a focus on engaging injured populations, represents a pragmatic approach to reach higher-risk persons already engaged in care.^18,31^

Kenya’s national HTS guidelines recommend HIV screening and targeted testing during all health-facility interactions with the goal of testing quarterly for KPs and every 6-12 months for PPs.^8^ Although this includes EDs, there is no specific program guidance to inform ED-HTS delivery. To address this, data from patient and healthcare provider stakeholders from the Kenyatta National Hospital (KNH) ED, in Nairobi, Kenya,^32,33^ were used to design the HIV Enhanced Access Testing in Emergency Departments (HEATED) program to improve ED-HTS delivery.

## METHODS

This quasi-experimental prospective study evaluated implementation of the HEATED program using the Reach, Effectiveness, Adoption, Implementation, and Maintenance (RE-AIM) framework applied to systems, patient and healthcare provider level data.^34^ Study periods included: pre-implementation (6 March – 16 April 2023), implementation (17 April – 30 April 2023), post-implementation period 1 (1 May – 26 June 2023) and post-implementation period 2 (27 June – 20 August 2023). Guidelines on implementation studies were adhered to in reporting these results.^35^

### Study Setting

The KNH ED, in Nairobi, Kenya provides 24-hour care access. At study initiation, there were 143 nurse, and physician staff members and eight HIV-services personnel working in the clinical setting. The ED offers continuous HTS with dedicated space and staffing for services. HTS is provided free of charge, using Alere Determine™ HIV-1/2 assays or OraQuick Rapid™ HIV-1/2 HIV self-test (HIVST) kits. Standardized records for all patients undergoing HTS are maintained using national reporting procedures, and any PLH identified is provided free follow up care at a government facility. Kenya’s national guidelines stipulate that screening for, and delivery of HTS, should occur during facility-based healthcare encounters = and recommends testing frequencies of every two years for the general population, quarterly for KPs and every 6-12 months for PPs.^5,36^

### HIV Enhanced Access Testing in Emergency Departments Program

The HEATED program aims to increase ED-HTS delivery and improve engagement for underserved higher-risk persons by supporting HIV service for injured persons and KP/PPs. The program was designed to be a pragmatic multi-component intervention which employs setting appropriate micro-strategies focused on HTS sensitization and integration, task shifting, resource reorganization, linkage advocacy, skills development and education. The HEATED program was developed through qualitative research from healthcare providers and patient stakeholders from the KNH emergency care setting.^32,33^ Qualitative data were mapped to the Capability-Opportunity-Motivation Behavioral model for change and the Theoretical Domains Framework to identify data-driven micro-strategies to enhance ED-HTS.^37,38^ A multipronged implementation strategy including training and educating, changing infrastructure and developing stakeholder interrelationships was used to implement the HEATED program (Supplement 1).^39^

### Data Sources

#### Systems-Level Data

Systems level data were collected from standardized clinical sources. The ED triage register was used to identify numbers of persons treated during study periods. HTS registries provided data on facility-based testing, HIVST provision and follow up and identification and linkage to care for PLH. Data were abstracted by trained study personnel and into digital databases with continuous quality monitoring.^40^

Data were collected from healthcare personnel. ED-HIV services providers were screened, consented and enrolled as participants. They were sampled during each study period using the Continuing professional development (CPD) Reaction questionnaire oriented to the research topic of ED-HTS for injured and higher-risk persons.^41^ The CPD-Reaction questionnaire is a validated instrument used in implementation sciences research, for assessing the impact of professional development activities on clinical behaviors.^42,43^ Anonymized data were collected during post-implementation period 2 from ED nurses and physicians via open ended self-completed surveys on perceptions of the HEATED program.

#### Patient Participant-Level Data

During pre-implementation and post-implementation period 1, data were obtained from enrolled patient participants. Trained study personnel present in the ED 24-hours a day collected standardized survey data. All patients ≥18 years, seeking injury care were eligible for participation. Injury designation used the standardized triage classification in the study setting.^44^ Patients known to be pregnant, incarcerated persons and those unable or unwilling to provide consent were excluded.

After provision of written informed consent, patient participants had enrollment data collected as close to ED arrival as possible on sociodemographics, past medical and social histories, HIV risk factors including screening for KPs and PPs. PPs based on Kenya’s national guidelines included young persons (18-24 years), victims of interpersonal violence (IPV), persons never previously HIV tested and persons with hazardous alcohol use (HAU).^5^ The Alcohol Use Disorders Identification Test (AUDIT-C) was used to identify HAU.^45,46^ At emergency care completion, patient participant data were collected on awareness and understanding of ED-HTS and services received. For patient participants who received ED-HTS, systems level data were linked and cross-referenced to validate report.

### Evaluation Approach

HEATED program implementation was assessed using the RE-AIM framework.^34^ The evaluation metrics for each dimension are shown in Table 1. Reach, effectiveness and implementation, data were assessed using systems and patient participant data. For adoption and maintenance, systems level data were used. CPD-Reaction scores from ED-HIV service providers were compared pre-implementation and post-implementation period 1. The constructs of intention and social influence, which represent willingness by the primary intervention agent to initiate a health behavior, were used for adoption. Constructs of beliefs about capabilities and moral norms representing the intervention agents’ fidelity to deliver the health service were used in assessment of implementation. Final CPD-Reaction scores obtained during post-implementation period 2, were used in assessment of maintenance, along with the longitudinal systems-level trends in HTS delivery

**Table 1.**
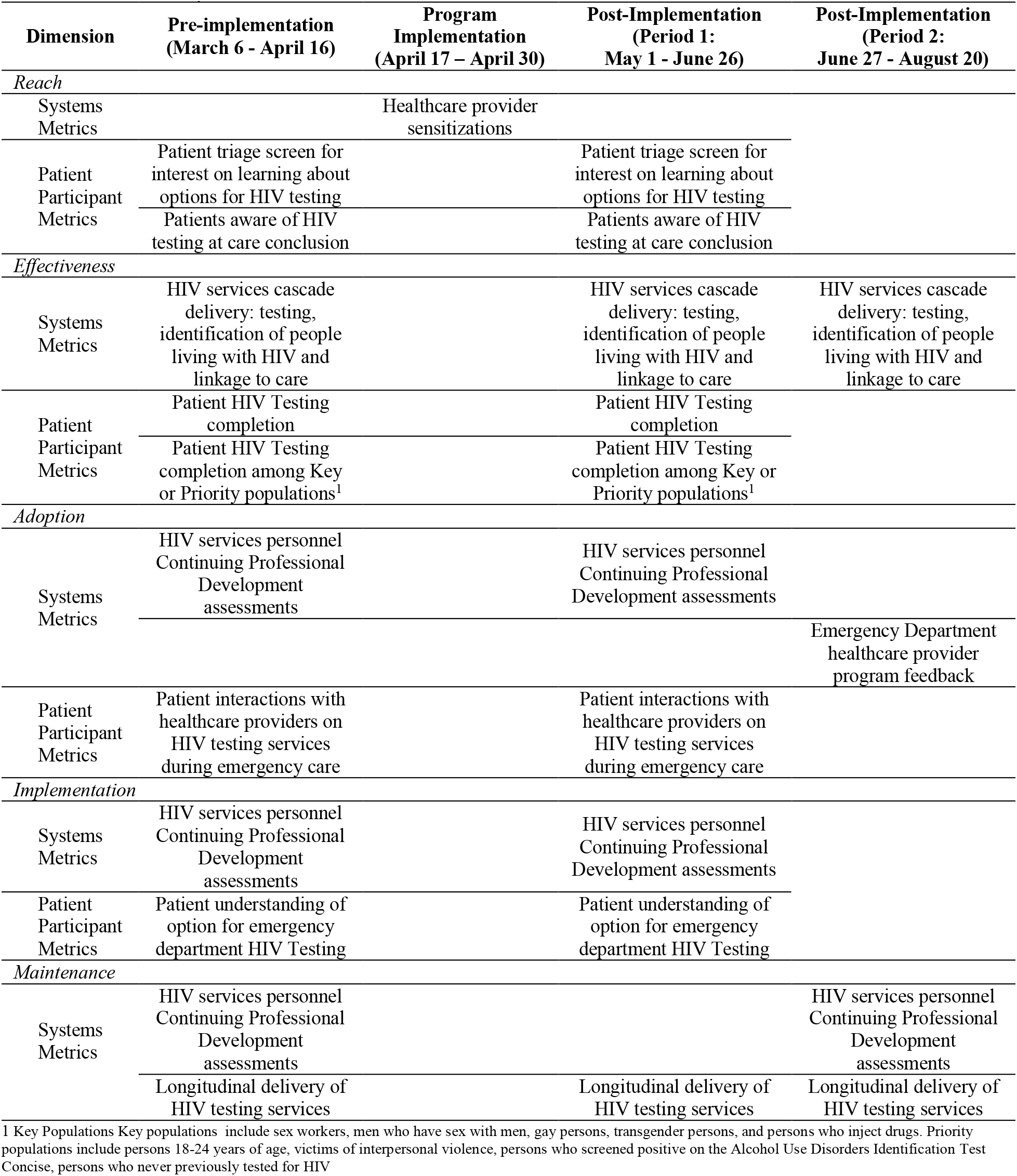
HIV Enhanced Access Testing in the Emergency Department Program Evaluation Assessment Metrics Across Study Periods.

### Statistical Analysis

The sample size was based on the outcome of increasing the proportion HIV test completed for injured persons seeking emergency care among enrolled patient participants. Prior data from the study setting demonstrated that 5.6% of injured patients completed ED-HIV testing.^25^ To be able to identify a 10% absolute increase in the proportion of injured patients completing testing, a sample size of ≥175 participant per assessment period was need (80% power, alpha <0.05).^47^

Descriptive analyses were performed for systems level and participant level data using frequencies with percentages or medians with associated interquartile ranges (IQR) as appropriate. Level of agreement for 5-point scale Likert scale items were summarized using medians with IQRs. HTS data were evaluated as facility-based HIV testing and distribution of HIVSTs independently, and aggregated as ED-HTS. Moving biweekly averages for HTS with associated variance estimates were calculated and graphed over time. Comparative analyses based on study period were performed using Pearson *X*^2^ or Fisher’s exact tests and Mann-Whitney tests as appropriate. Using the pre-implementation data as the baseline comparator, risk ratios (RR) were calculated with associated 95% confidence intervals (CI) for HTS cascade metrics for systems level and participant level data. An *a priori* subgroup analysis was completed on HIV testing for KPs and PPs among enrolled patient participants.

### Ethical Approvals

The study was approved by the University of Nairobi ethics and research committee and the institutional review board of Rhode Island Hospital. All enrolled participants provided written informed consent.

## RESULTS

### Study Population

#### Systems-level

There were a total of 12,532 ED encounters during data collection periods. Of these, 3,050 (24.4%) received ED-HTS, with 696 pre-implementation and 2,354 post-implementation (Figure 1 Panel A). The majority of persons who received facility-based HTS were male and >25 years of age. Across all periods, approximately two-thirds of persons completing facility-based HTS had never previously been tested for HIV (Table 2).

**Figure 1:**
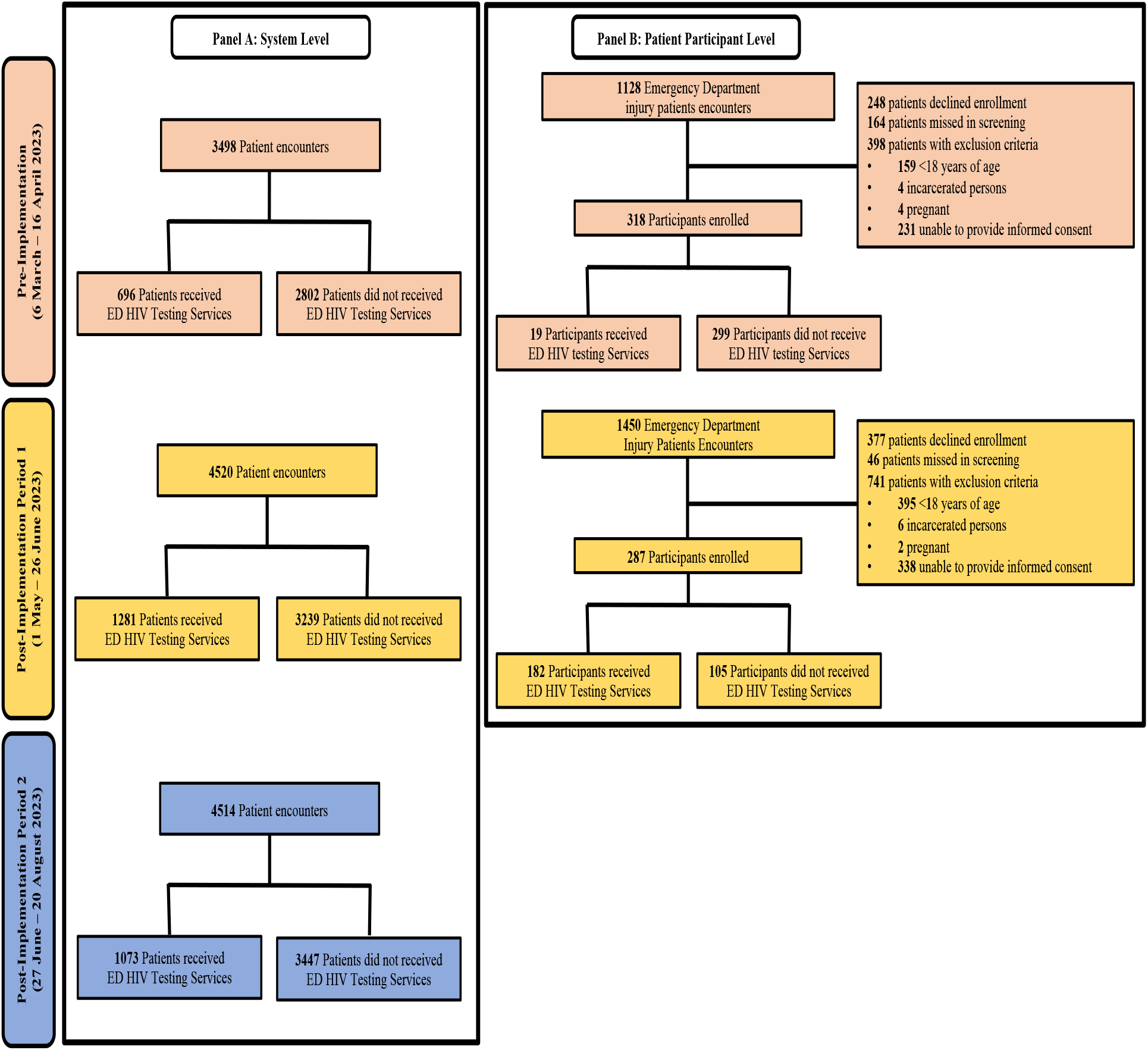
Study Population

**Table 2.** System-level Data on Recipients of HIV Testing Services.

|  | **Pre-**<br>**implementation**<br>**(March 6 –**<br>**April 16)**<br>**n (%)** | **Post-**<br>**Implementation**<br>**(Period 1: May 1**<br>**- June 26)**<br>**n (%)** | **p-**<br>**value** | **Post-**<br>**Implementation**<br>**(Period 2: June**<br>**27 - August 20)**<br>**n (%)** | **p-**<br>**value** |
| --- | --- | --- | --- | --- | --- |
| Completed Facilities-based HIV Testing | 585 | 1031 |  | 860 |  |
| Gender |  |  |  |  |  |
| Male | 296 (50.6) | 574 (55.7) | 0.046 | 503 (58.5) | 0.008 |
| Female | 286 (48.9) | 456 (44.2) |  | 350 (40.7) |  |
| Transgender | 0 (0.0) | 0 (0.0) |  | 0 (0.0) |  |
| Missing | 3 (0.6) | 1 (0.1) |  | 7 (0.8) |  |
| Age years (median) | 34 (25, 43) | 35 (27, 44) | 0.313 | 33 (26, 43) | 0.775 |
| Age 15-24 years | 95 (16.2) | 164 (15.9) | 0.861 | 159 (18.5) | 0.270 |
| Key Population<sup>1</sup> | 1 (0.1) | 3 (0.2) | 1.00 | 10 (1.2) | 0.058 |
| Ever previously Tested for HIV |  |  |  |  |  |
| No | 381 (65.1) | 673 (65.3) | 0.046 | 565 (65.7) | 0.027 |
| Yes | 194 (33.2) | 353 (34.2) |  | 292 (36.0) |  |
| Missing | 10 (1.7) | 5 (0.5) |  | 3 (0.03) |  |
| Used HIV self-test in last 12 months |  |  |  |  |  |
| No | 571 (97.6) | 1020 (98.93) | 0.038 | 857 (99.7) | <0.001 |
| Yes | 14 (2.4) | 11 (1.07) |  | 3 (0.3) |  |
| Persons accepting HIV self-test kits | 111 | 250 |  | 213 |  |
| Gender |  |  |  |  |  |
| Male | 49 (44.1) | 128 (51.2) | 0.090 | 138 (64.8) | <0.001 |
| Female | 59 (53.2) | 121 (48.4) |  | 74 (34.7) |  |
| Transgender | 0 (0.0) | 0 (0.0) |  | 0 (0.0) |  |
| Missing | 3 (2.7) | 1 (0.4) |  | 1 (0.5) |  |
| Age 15-24 years | 33 (29.7) | 38 (15.2) | 0.084 | 49 (23.0) | 0.186 |

<sup>1</sup>Key populations include sex workers, men who have sex with men, gay persons, transgender persons and persons who inject drugs.

#### Patient Participant level

During pre-implementation and post-implementation period 1, there were 2,578 injury encounters. Of these, 2,303 (89.3%) were screened, 605 (26.3%) met inclusion and were enrolled. During pre-implementation 19 of 318 participants received ED-HTS; and 182 of 287 participants received ED-HTS post-implementation (Figure 1, Panel B). Patient participant characteristics were similar between pre-implementation and post-implementation period 1. The majority of participants were male and approximately one-quarter were <25 years of age. KPs comprised 5.0% of patient participants pre-implementation and 6.3% in post-implementation period 1 (p=0.508). PPs were 67.0% of participants pre-implementation and 63.3% in post-implementation period 1 (p=0.640). Similar portions of participants had never been HIV tested 12.9% pre-implementation versus 12.6% post-implementation (p=0.881). One third of participants had a history of interpersonal violence (33.3% pre-implementation, 33.1% post-implementation, p=0.952) (Table 3).

**Table 3.** Patient Participant Characteristics.

|  | **Pre-implementation**<br>**(March 6 - April 16)** | **Post-Implementation**<br>**(Period 1: May 1 - June 26)** | **p-value** |
| --- | --- | --- | --- |
|  | **n (%)** | **n (%)** |  |
| Age (median, IQR) | 31 (25-38) | 30 (24-38) | 0.655 |
| Young adult (18-24 years) | 76 (23.9) | 76 (26.6) | 0.450 |
| Gender |  |  |  |
| Male | 263 (82.7) | 231 (80.5) |  |
| Female | 53 (16.7) | 54 (18.9) | 0.608 |
| Transgender | 1 (0.3) | 0 (0.0) |  |
| Missing | 0 (0.0) | 2 (0.7) |  |
| Relationship Status |  |  |  |
| Single | 115 (36.3) | 138 (48.3) |  |
| Married | 163 (51.4) | 96 (33.2) |  |
| In a monogamous relationship | 17 (5.4) | 32 (11.2) |  |
| In a polygamous relationship | 1 (0.3) | 4 (1.4) | <0.001 |
| Separated (still married) | 16 (5.1) | 10 (3.5) |  |
| Wishes not to disclose | 5 (1.6) | 5 (1.8) |  |
| Missing | 1 (0.3) | 2 (0.7) |  |
| Education |  |  |  |
| Primary schooling or less | 91 (28.6) | 97 (33.8) |  |
| Secondary schooling or greater | 225 (70.8) | 188 (65.5) | 0.328 |
| Missing | 2 (0.6) | 2 (0.7) |  |
| Employment Status |  |  |  |
| Employed | 236 (74.2) | 233 (81.2) |  |
| Unemployed | 77 (24.2) | 52 (18.1) | 0.102 |
| Missing | 5 (1.6) | 2 (0.7) |  |
| Has Established Primary Care Provider |  |  |  |
| No | 260 (81.8) | 234 (81.5) |  |
| Yes | 56 (17.6) | 51 (17.8) | 0.992 |
| Missing | 2 (0.6) | 2 (0.7) |  |
| Identifies as a Key Population<sup>1</sup> | 16 (5.0) | 18 (6.3) | 0.508 |
| Characterized as a Priority Population<sup>2</sup> | 213 (67.0%) | 182 (63.4%) | 0.671 |
| Never Previously Tested for HIV | 41 (12.9) | 36 (12.6) | 0.881 |
| Hazardous Alcohol Use<sup>1</sup> | 80 (25.2) | 70 (24.5) | 0.827 |
| History of Interpersonal Violence |  |  |  |
| No | 212 (66.7) | 192 (66.9) |  |
| Yes | 106 (33.3) | 95 (33.1) | 0.952 |
| Condomless sex in last six months |  |  |  |
| No | 164 (51.6) | 157 (54.6) |  |
| Yes | 142 (44.7) | 117 (40.9) | 0.904 |
| missing | 2 (0.6) | 2 (0.7) |  |
| Emergency Department Disposition |  |  |  |
| Discharged | 175 (55.0) | 36 (60.0) |  |
| Admitted | 130 (41.0) | 23 (38.3) |  |
| Eloped | 6 (1.9) | 0 (0.0) | 0.681 |
| Missing | 7 (2.2) | 1 (1.67) |  |

<sup>1</sup> Key Populations Key populations include sex workers, men who have sex with men, gay persons, transgender persons, and persons who inject drugs
<sup>2</sup> Priority populations include persons 18-24 years of age, victims of interpersonal violence, persons who screened positive on the Alcohol Use Disorders Identification Test, persons who never previously tested for HIV

### Reach

All 151 ED personnel were reached though HEATED program sensitization. In-person sessions were attended by 45% of personnel, and all personnel received digital communications. All eight ED-HIV services providers completed HEATED program training on HTS for underserved higher-risk persons (Supplement 1). Among patient participants, screening at triage on interest in learning about ED-HTS options increased from 3.5% pre-implementation to 46.7% post-implementation period 1 (p<0.001). At ED care completion, patient participant awareness of ED-HTS options increased from 37.7% pre-implementation 76.9% to post-implementation period 1 (p<0.001) (Table 5).

### Effectiveness

ED-HTS, which aggregated facility-based HIV testing and distribution of HIVSTs, increased from 19.9% (95% CI: 18.6-21.3) pre-implementation to 28.5% (95% CI: 27.2–29.8) post-implementation period 1 and 23.8 (95% CI: 22.6-25.1) in post-implementation period 2. As compared to pre-implementation, systems level ED-HTS was significantly greater in the post-implementation periods (RR=1.31, 95% CI:1.21-1.43; p<0.001) (Figure 2 Panel A). Identification of PLH non-significantly increased from 24 (4.1%) pre-implementation to 53 (5.1%) post-implementation period 1 and to 43 (5.0%) post-implementation period 2. Among those tested in the ED and found to be PLH, 73% were newly diagnosed. Linkage to care of eligible PLH was 71.4.% pre-implementation, 81.1% in post-implementation period 1 (p=0.454) and 87.0% in post-implementation period 2 (p=0.242). The number of persons distributed HIVST kits increased with program implementation and there was a significant increase in follow up of usage by ED-HIV services providers (Table 4).

**Figure 2.**
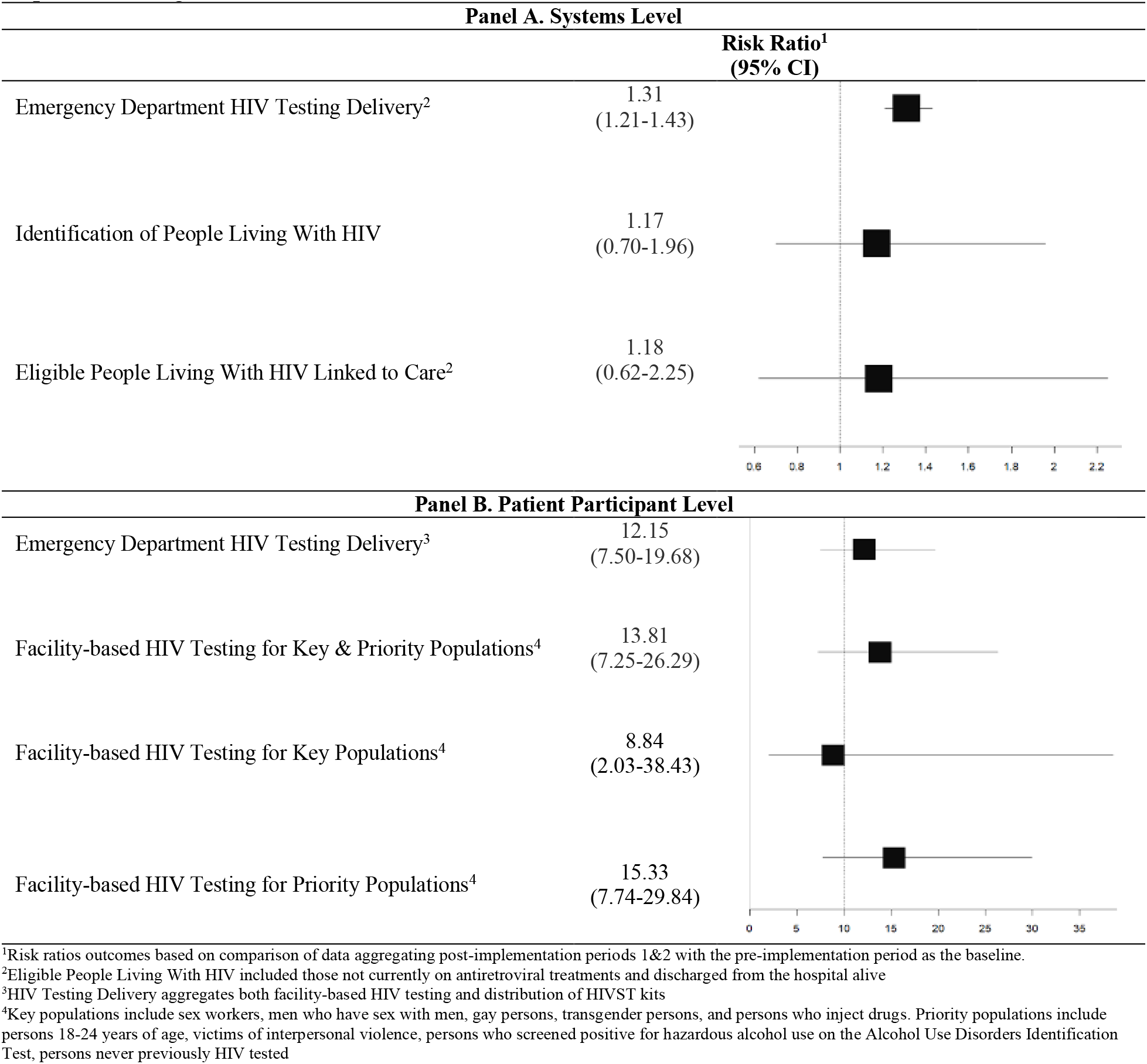
HIV Services With Implementation of the HIV Enhanced Access Testing in the Emergency Department Program

**Table 4.** System Level Emergency Department HIV Testing Services.

|  | Pre-implementation<br>(March 6 - April<br>16)<br>n (%) | Post-Implementation<br>(Period 1: May<br>1 - June 26)<br>n (%) | p-value | Post-Implementation<br>(Period 2: Jun<br>27 - August 20)<br>n (%) | p-value |
| --- | --- | --- | --- | --- | --- |
| Facility-based HIV testing | 585 | 1031 |  | 860 |  |
| HIV Test Result |  |  |  |  |  |
| Negative | 560 (95.7) | 977 (94.8) | 0.453 | 816 (94.9) | 0.703 |
| Positive | 24 (4.1) | 53 (5.1) |  | 43 (5.0) |  |
| Inconclusive | 1 (0.2) | 1 (0.1) |  | 1 (0.1) |  |
| Positive HIV Test Results |  |  |  |  |  |
| Previously diagnosed | 8 (33.3) | 13 (24.5) | 0.555 | 15 (34.9) | 0.239 |
| New Diagnosis | 16 (66.7) | 40 (75.5) |  | 28 (65.1) |  |
| New Diagnosis Linked to Care |  |  |  |  |  |
| No (declined) | 4 (25.0) | 7 (17.5) | 0.657 | 3 (10.7) | 0.447 |
| No (deceased) | 2 (12.5) | 3 (7.5) |  | 5 (17.9) |  |
| Yes | 10 (62.5) | 30 (75.0) |  | 20 (71.4) |  |
| New Eligible Diagnosis Linked to Care |  |  |  |  |  |
| No (declined) | 4 (28.6) | 7 (18.9) | 0.454 | 3 (13.0) | 0.242 |
| Yes | 10 (71.4) | 30 (81.1) |  | 20 (87.0) |  |
| Previous Diagnosis Linked to Care |  |  |  |  |  |
| No (Already linked to care) | 7 (87.5) | 8 (61.5) | 0.201 | 10 (66.7) | 0.651 |
| No (Declined) | 0 (0.0) | 0 (0.0) |  | 2 (13.3) |  |
| Yes | 1 (12.5) | 5 (38.4) |  | 3 (20.0) |  |
| Previous Diagnosis Linked to Care (Eligible) |  |  |  |  |  |
| No (Declined) | 0 (0.0) | 0 (0.0) | - | 2 (40.0) | 0.439 |
| Yes | 1(100.0) | 5 (100.0) |  | 3 (60.0) |  |
| Provision of HIV Self-Test |  |  |  |  |  |
| HIV Self-Test Recipients | 111 | 250 |  | 213 |  |
| HIV Self-Test Follow-up Completed |  |  |  |  |  |
| No | 21 (18.92) | 64 (25.6) | <0.001 | 42 (19.7) | <0.001 |
| Yes | 0 (0.0) | 186 (74.4) |  | 171 (80.3) |  |
| Missing | 90 (81.09) | 0 (0.0) |  | 0 (0.0) |  |
| HIV Self-Test Reported Use |  |  |  |  |  |
| No | - | 10 (5.4) | - | 10 (5.9) | - |
| Yes | - | 176 (94.6) |  | 161 (94.2) |  |
| HIV Self-Test Reported Result |  |  |  |  |  |
| Non-reactive | - | 176 (100.0) | - | 159 (98.8) | - |
| Reactive | - | 0(0.0) |  | 1(0.6) |  |
| Inconclusive | - | 0(0.0) |  | 1(0.6) |  |

ED-HTS significantly increased among patient participants from 5.7% pre-implementation to 69.0% post-implementation period 1. There were three patient participants identified as PLH. Figure 3 shows changes in HTS cascade delivery with HEATED program implementation stratified by participants as KPs/PPs or not. For KPs/PPs participants facility-based HIV testing increased from 4.6% pre-implementation to 72.3% post-implementation period 2 (p<0.001). Among KPs/PPs completing facility-based HIV testing, 49% had multiple KP identifies and/or multiple categorizing PP characteristics (Table 5). As compared to pre-implementation, patient participants in post-implementation period 1 were significantly more likely to complete ED-HTS (RR=12.15, 95% CI:7.50-19.68; p<0.001). This significant increase was also found in the KPs (RR=8.84, 95% CI:2.03-38.43; p<0.001) and the PP (RR=15.22, 95% CI:7.74-29.94; p<0.001) (Figure 2 Panel B).

**Figure 3.**
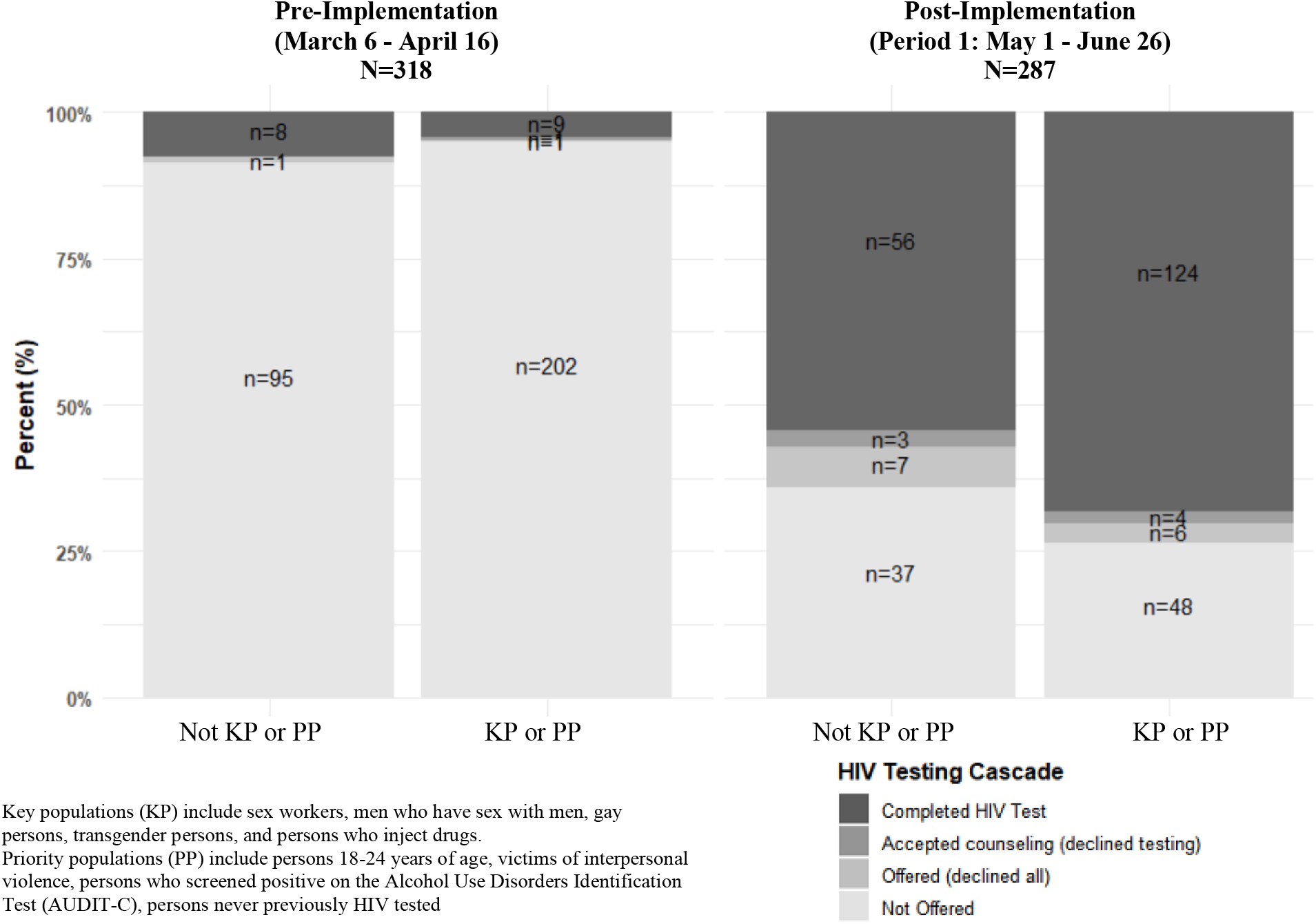
HIV Testing Cascade Pre- and Post-implementation of the HIV Enhanced Access Testing in the Emergency Department Program Stratified by Key and Priority Populations

**Table 5.** HIV Testing Services For Patient Participants From Key and Priority Populations.

|  | Pre-implementation<br>(March 6 - April<br>16)<br>N=216<br>n (%) | Post-Implementation<br>(Period 1: May 1 -<br>June 26)<br>N=184<br>n (%) | p-value |
| --- | --- | --- | --- |
| Key & Priority Populations<sup>1</sup> |  |  |  |
| Not Receiving HIV Testing Services | 206 (95.4) | 51 (27.7) | <0.001 |
| Receiving HIV Testing Services | 10 (4.6) | 133 (72.3) |  |
| Key & Priority Populations<sup>1</sup> Receiving HTS |  |  |  |
| Young Adult | 1 (10.0) | 14 (14.7) |  |
| & Never Previously Tested for HIV | 1 (10.0) | 10 (7.5) |  |
| & Hazardous Alcohol Use | 1 (10.0) | 1 (0.8) |  |
| & Interpersonal Violence Victim | 1 (10.0) | 11 (8.3) |  |
| & Sex Work | 0 (0.0) | 2 (1.5) |  |
| & Never Previously Tested for HIV & Interpersonal Violence<br>Victim | 0 (0.0) | 4 (3.0) |  |
| & Interpersonal Violence Victim & Hazardous Alcohol Use | 0 (0.0) | 3 (2.3) |  |
| & Interpersonal Violence Victim & Sex Work | 0 (0.0) | 2 (1.5) |  |
| & Never Previously Tested for HIV & Sex Work & Gay | 0 (0.0) | 1 (0.8) |  |
| & Interpersonal Violence Victim & Sex Work<br>& Gay | 0 (0.0) | 1 (0.8) |  |
| Never Previously Tested for HIV | 0 (0.0) | 4 (3.0) |  |
| & Hazardous Alcohol Use | 1 (10.0) | 4 (3.0) |  |
| & Interpersonal Violence Victim | 0 (0.0) | 1 (0.8) |  |
| & Sex Work | 0 (0.0) | 1 (0.8) |  |
| Hazardous Alcohol Use | 1 (10.0) | 20 (15.0) |  |
| & Interpersonal Violence Victim | 0 (0.0) | 15 (11.3) |  |
| & Sex Work | 0 (0.0) | 4 (3.0) |  |
| & Gay | 1 (10.0) | 0 (0.0) |  |
| & Interpersonal Violence Victim & Sex Work | 0 (0.0) | 2 (0.8) |  |
| & Interpersonal Violence Victim & Person Who Injects Drugs | 0 (0.0) | 1 (0.8) |  |
| & Interpersonal Violence Victim & Sex Work<br>& Person Who Injects Drugs | 0 (0.0) | 1 (0.8) |  |
| Interpersonal Violence Victim | 2 (20.0) | 30 (22.6) |  |
| Sex Work | 0 (0.0) | 1 (0.8) |  |
| Gay | 1 (10.0) | 0 (0.0) |  |

<sup>1</sup> Key Populations Key populations include sex workers, men who have sex with men, gay persons, transgender persons, and persons who inject drugs. Priority populations include young adult persons 18-24 years of age, victims of interpersonal violence, persons who screened positive on the Alcohol Use Disorders Identification Test for Hazardous Alcohol use, persons who never previously tested for HIV

### Adoption

ED-HIV services personnel CPD-Reactions scores increased pre-implementation period to post-implementation period 1. For the construct of intention median scores increased from 6.3 (IQR: 5,8, 6.8) to 6.5 (IQR: 6.5, 7.0) (p=0.682) and from 4.3 (IQR: 3.4, 4.7) to 5.2 (IQR: 4.3, 5.8) (p=0.014) for social influence (Supplement 2). Anonymous feedback from ED healthcare personnel was mixed with some personnel indicating that the HEATED program improved testing and was used well. However, there was concerns noted that not all staff were engaged (Supplement 3).

### Implementation

Among ED-HIV services personnel scores for CPD-Reaction constructs of beliefs about capabilities and moral norms significantly increased. Median scores increased from 5.2 (IQR: 4.7, 5.7) pre-implementation to 6.0 (IQR: 5.8, 6.3) post-implementation period 1 (p=0.013) for beliefs about capabilities. For moral norms, median scores increased pre-implementation from 6.0 (IQR: 5.5, 6.0) to 6.8 (IQR: 6.0, 7.0) post-implementation period 1 (p=0.014) (Supplement 2). During pre-implementation 3.8% of enrolled patient participants reported discussing HTS with ED personnel during care, which increased to 85.3% post-implementation (p<0.001). There was a significant increase in agreement among enrolled patient participants that it was clear that ED-HTS was available with median Likert scores increasing from 2 (IQR: 1, 5) pre-implementation to 5 (IQR: 2, 5) post-implementation period 1 (P<0.001)

### Maintenance

In post-implementation periods 1 and 2, encompassing 16 weeks of systems level data, a significant increase in ED-HTS delivery was maintained as compared to pre-implementation. Figure 4 shows bi-weekly average proportional ED-HTS delivery. Facility-based HTS ranged from 13.9% to 19.1% during pre-implementation and from 17.1% to 25.4% post-implementation. Bi-weekly average HIVST distribution ranged from 1.6% to 3.7% pre-implementation and from 2.5% to 8.4% post-implementation. CPD-Reaction scores for ED-HIV services personnel were persistently greater on follow up assessments during post-implementation period 2 as compared to pre-implementation (Supplement 2).

**Figure 4.**
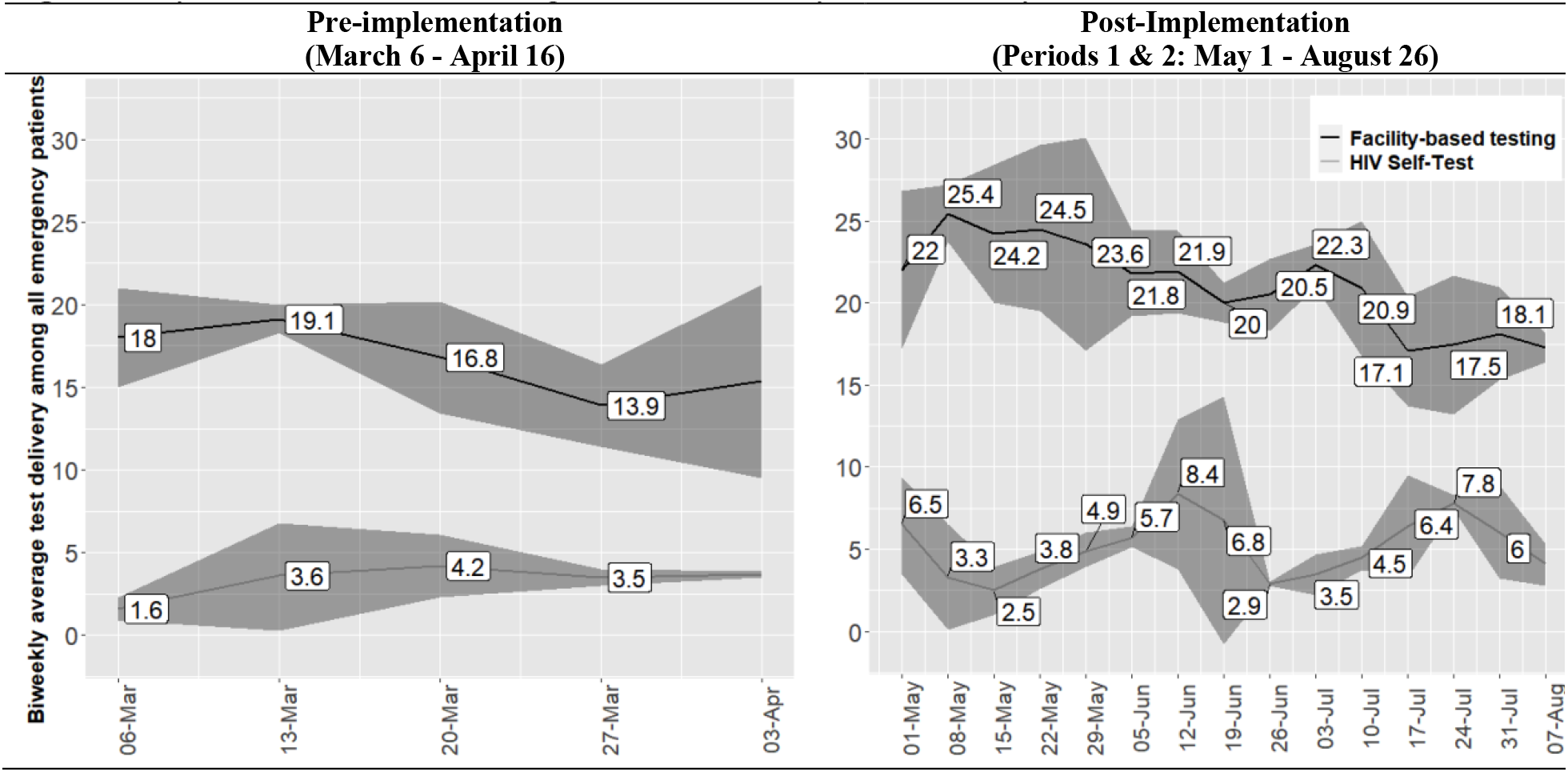
Systems Level HIV Testing Services Delivery Across Study Periods^1^

## DISCUSSION

The current study assessing implementation of the HEATED program demonstrates positive impacts on ED-HTS across the RE-AIM domains. The systems-level and patient participant data showed significantly increased testing services, suggesting that broader program development may represent a pragmatic approach to augment HTS delivery to higher-risk target populations. Considering the burdens of injury across sub-Saharan Africa requiring emergency care,^26,27^ particularly in underserved KP/PPs,^28,29^ and barriers to reaching those persons for HIV services,^14–16^ further evaluation of the HEATED program and leveraging of ED-HTS as a mechanism to contribute to reaching global HIV control targets is warranted.

The HEATED program used multi-modal approaches with in person trainings, digital contact for sensitization and one-on-one information delivery to reach all providers in the study setting. However, feedback indicated greater sensitization was needed, suggesting that program reach could have been improved. Although, more sensitization may have been beneficial, the significant increase in ED-HTS delivery with program implementation suggests that willingness to improve services was achieved. This is also supported by patient data showing a significant increase in triage screening for interest in learning about ED-HTS options during emergency care. As triage was performed by varying staff, the significant increase in screening suggests that the HEATED program reached intended providers. However, as ensuring all stakeholders are reached in an acceptable and consistent manner is crucial to HIV program delivery,^48^ and given changes in staffing and policies that occur over time, it is likely the HEATED program could require more intensive and longitudinal sensitization to sustain gains in HTS impacts.

The primary outcome of effectiveness to improve ED-HTS among injured patient participants demonstrated that the HEATED program was associated with a significant increase in testing. Within the enrolled sample, one in every 18 patients was from a KP, and the majority of participants had characteristics for PPs. These data have external validity, with injury profiles from sub-Saharan Africa demonstrating high burdens among KPs,^28,29^ and the epidemiology of those most likely to suffer injuries being men, young people and persons who use drugs,^26,27,49^ supporting the importance of the ED setting for delivery of HTS. Additionally, as nearly half of enrolled patient participants had multiple KP identities and/or PP characteristics, and prior data has shown greater HIV vulnerability among such persons,^50^ the ED may represent an important venue to reach individuals with compounding HIV risk. Although, there was a low number of PLH identified among the patient participants, the underserved population reached through the injury focus highlights the potential for ED-HTS to pragmatically deliver services to targeted groups already in contact with healthcare. The HEATED program approach is consistent with recommendations for HIV programming for KPs, men and young adults, in improving access to services by meeting persons where they are and reducing gender barriers and stigmatization by utilizing the lens of injury which is not dependent on population identity.^10,12,13^ As patient ED-HTS acceptability in sub-Saharan Africa is high,^23–25,51,52^ further study of how to ensure systems support healthcare providers to best achieve delivery will be beneficial to build from the HEATED program results.

Effectiveness in ED-HTS was reproduced in the systems-level data with significant increases in the post-implementation periods. The increase in overall testing suggests that the HEATED program, did not redirect HTS, but rather increased services. This is evident in the observed increase in testing of men, who made up approximately 80% of injury patients in the study setting. Considering the majority of those injured globally are men,^26,53^ and that men in sub-Saharan Africa are inadequately tested, diagnosed later and have higher HIV-related mortality than females,^13,54^ the ED represents a venue deliver person-centered care in a gender-neutral setting to men, consistent with WHO recommendations.^13^ In the systems level data, there was a non-significant increase in PLH identified and linked to care following HEATED program implementation. The current study was not designed to assess post-testing HIV care cascade outcomes, and albeit suggestive of positive down-stream impacts of the HEATED program this is hypothesis generating, and a sufficiently powered interventional trial is needed to robustly assess this. Although achieving the UNAIDS 95-95-95 targets requires identification and linkage to treatment of PLH, status-neutral programming with interval testing and delivery of preventive interventions is also integral in epidemic control.^2,55,56^ As Kenya’s guidelines recommend HIV testing quarterly for KPs and every 6-12 months for PPs,^8^ further development of ED-HTS is not only important in completing the request first step of testing required for antiretroviral therapy (ART) or Pre-exposure Prophylaxis (PrEP) provision, but would also support achieving delivery of interval testing to targeted populations.

Correlating with the significant increase in systems-level HTS, assessments of the primary intervention agents, ED-HIV services personnel, found sustained increases in assessments for constructs on program adoption and implementation. Patient participant data showed program implementation fidelity for HTS sensitization, with significantly more patients aware of services and discussing testing with ED healthcare providers in the post-implementation period. However, the linked systems-level and patient participant level data demonstrated that only 12% of persons from KPs were identified during their receipt of ED-HTS. As a component of the HEATED program was training ED-HIV services personnel on interacting with and delivering care for KPs, this suggests that the program did not sufficiently meet the provider needs and research to inform approaches to improve care for KPs from emergency care settings is needed.

With the quasi-experimental design, causality cannot be determined. Generalizability of the HEATED program in alternative settings with differing systems and resources is not known. However, as the program was designed to be pragmatic and adaptable to the = relevant needs of a service delivery point it may be impactful in other settings, which requires further study. Due to the use of standard reporting tools from systems-level data, effects on preventative services such as risk reduction counseling, condom distribution and PrEP screening were not able to be assessed across study periods. Although, the RE-AIM framework was used, qualitative data focusing on adoption and implementation would have supported a more comprehensive evaluation of the HEATED program. As well, longer-term maintenance data on sustainability, costing analyses and outcomes on linkage to treatment would strengthen the application of program. To address this, and more robustly evaluate the HEATED program, a cluster randomized trial using mixed methods with a sufficient maintenance period and costing assessment is needed.

## CONCLUSIONS

The HEATED program significantly increased HTS delivery and augmented testing to underserved populations seeking emergency care. The program assessment across RE-AIM framework domains showed favorable data suggesting that further implementation study could support pragmatic service improvements for higher-risk persons already engaged with health systems as a mechanism to support advancement towards HIV control targets.

## Competing interests and sources of funding

None of the authors conflicts of interest. The contents of this manuscript are solely the responsibility of the authors and do not necessarily represent the official views of affiliated institutions or funding bodies. ARA and the overall work was supported by the National Institute of Allergy and Infectious Diseases (grant number: K23AI145411). JSS was supported in part by the National Institute of Allergy and Infectious Diseases (grant number: R25AI140490). AO was supported in part by the National Institute of Drug Abuse (grant number: T32DA013911). DAK was supported in part by University of Washington/Fred Hutch Center for AIDS Research, an NIH-funded program (grant number: P30 AI027757).

## Data availability statement

Deidentified data that support the findings of this study are available from the corresponding author upon reasonable request.

## Author Contributions

ARA, WM, SP, JK, DB, HW, RB, DKO, DAK, MJM CF contributed to the conceptualization of the topic. ARA, JS, AO, WM, SP, JK, DB, HW, RB contributed to the data curation. ARA, JS, AO, HW analyzed the data and drafted the paper. Funding was acquired by ARA, JK, RB, DAK, MJM CF. All authors read, commented on and approved the final manuscript.

## Supplements

### Supplement 1. HIV Enhanced Access Testing in Emergency Departments Program

Based on prior data from the Kenyatta National Hospital (KNH) study setting demonstrating potential to improve emergency department (ED) HIV Testing services (HTS) delivery, particularly among injured persons seeking care, qualitative studies were undertaken with healthcare provider and patient stakeholders. The qualitative research aimed to evaluated challenges and facilitator for HTS programming within the KNH ED. The Capability-Opportunity-Motivation Behavioral Model (COM-B), which examines health behaviors through three interrelated lenses, was utilized as a framework to overlay the key findings of the thematic analysis, and the Theoretical Domains Framework (TDF) was used to identify intervention functions to inform approaches to enhance ED-HTS programming. Following the initial design phase feedback sessions with healthcare and administrative personnel from the KNH setting were held to review and refine the design. This step resulted in the addition of external training from the Kenya National AIDS and STIs Control Programme (NASCOP) pertaining to Key Populations (KPs) for the ED-HIV services personnel. The HEATED program components across the COM-B model are shown and described below. All aspects of HEATED program were initiated during the implementation period (17-30 April 2023), except for the change made to the standard triage process in which a screening question on interest in learning about HTS during ED care was added at the start of the pre-implementation data collection period (6 March 2023). The implementation strategy was based on Expert Recommendations for Implementing Change and was multipronged and included micro-stratgies aiming to change infrastructure, supporting clinicians, providing interactive assistance, train and educate stakeholders, develop stakeholder relationships, and engaging patients. This approach was used to provide an evidence-based program with components which were locally appropriate and feasible to be implemented.

Capability-Opportunity-Motivation Behavioral Model For the HIV Enhance Access Testing in Emergency Department Program

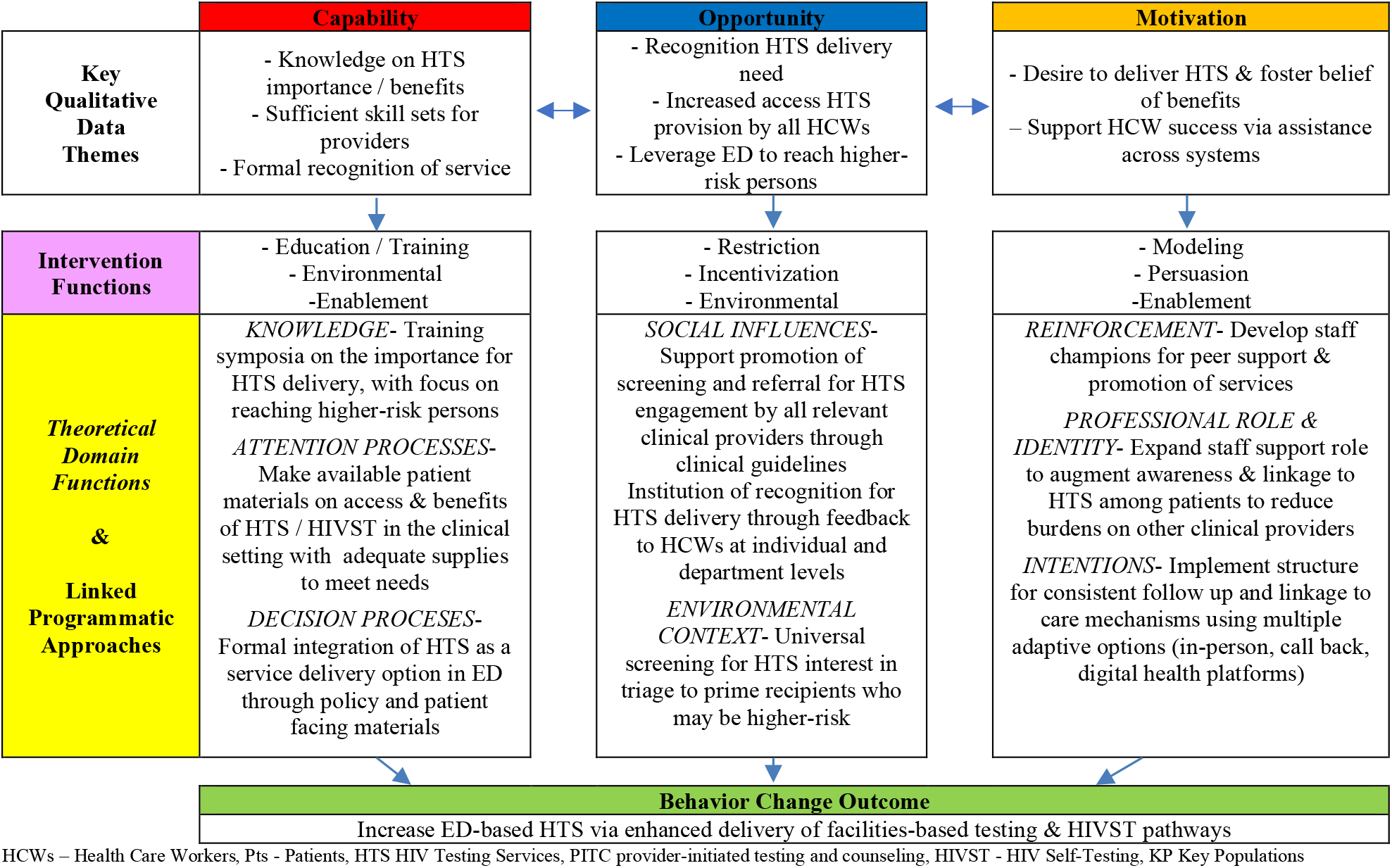

#### Capability

The domain of capability refers to an individual’s physical and psychological ability to engage in a health behavior of increasing HTS delivery with a focus on higher-risk persons in the ED setting. The key intervention functions for capability were education and training, environmental restructuring and enablement. Components for the HEATED program for Capability were:

##### Sensitization symposia

These training sensitized and provided educational information on the role and importance of HIV services in the ED setting and were held with staff inclusive of nurses, physicians, administrators and HIV services personnel. The content used NASCOP materials on HTS recommendations and policies as well as data from the study setting focusing on the local HTS landscape and how there exists opportunities to engage higher-risk persons for HIV services during emergency care, especially for injured persons. The sessions also provided an overview of the overall HEATED program and its components. These symposia were completed through multiple sessions and across varying venues in the ED. There were two sessions lasting 90 minutes in duration held through standard ED meeting mechanisms for continuing education and training, which were approved for professional development credit via KNH procedures. To help ensure support sensitization text messages on the program were sent to provider groups of nurses and physicians using their established communication list-serves with the ability to engage in discussion and clarification as needed with the healthcare champions who were nurse and physician members of the ED-staff. Additionally, four brief (15 minute) sessions were given during meetings with clinical staff in the ED space. The content was delivered by a combination of study team members and local HTS training personnel. During all sessions there was time allocated for questions and open-forum discussion.

##### HIV Services Personnel Engagement Training

Based on the observed low engagement of injured persons during ED care and the overlap of this population with higher-risk groups (i.e. KPs, young adults and men) a focused training on engaging such persons was provided to the HIV services personnel. The training utilized existing materials and facilitators from NASCOP which are designed to train up healthcare providers around engagement of target persons for HIV testing and care. Within this training there was also dedicated sessions on engaging the ED injury population for HTS. The training sessions were be held over two three-day periods to accommodate flexible attending for all HTS personnel.

##### Targeted advertising

Prior to program implementation in the ED there existed minimal signage and written information on the availability of HTS. Using NASCOP resources awareness material in the form of signs and educational pamphlets were placed in six locations in the ED at strategic points to enhance patient knowledge on HTS.

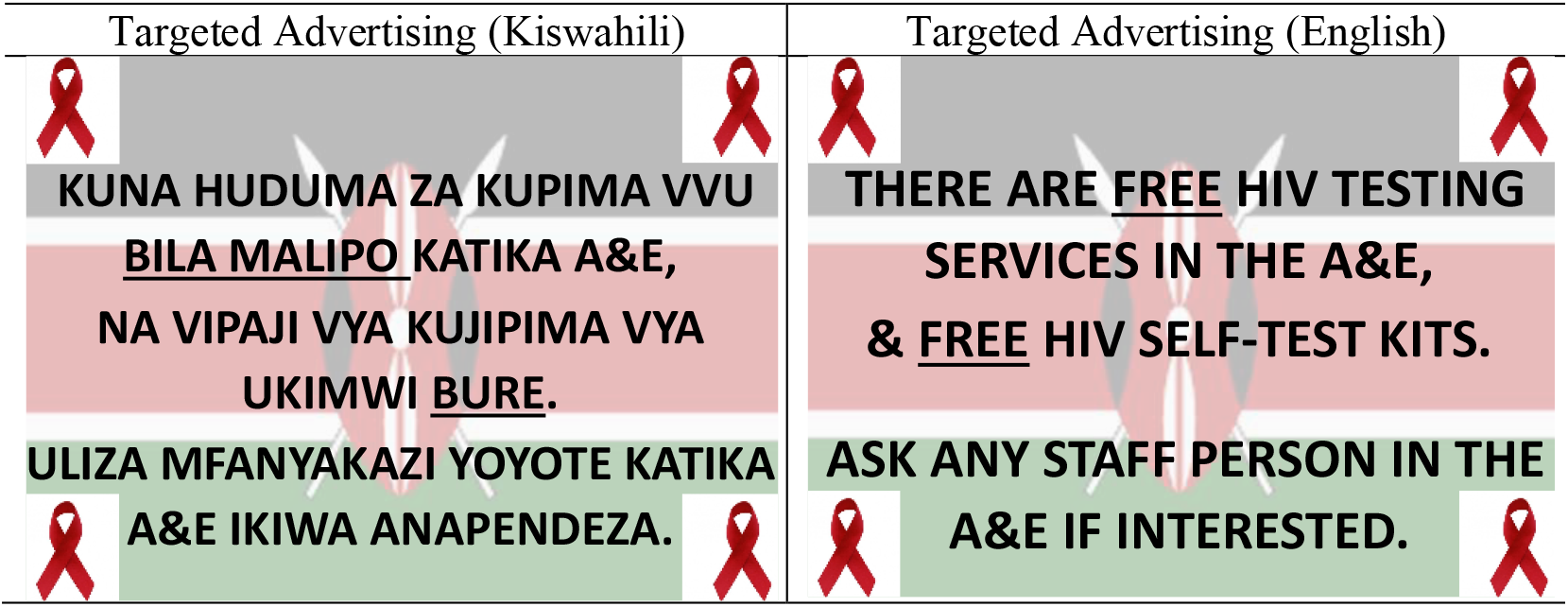

##### HTS Integration

Although the HTS services existed within the KNH ED space and care delivery system lack of coherent services integration was identified as a challenge to HTS. To address this the three ED flow maps, which provide a throughput overview of a patients’ ED care, were revised to include HTS (see figure). These replaced the existing versions in the clinical space.

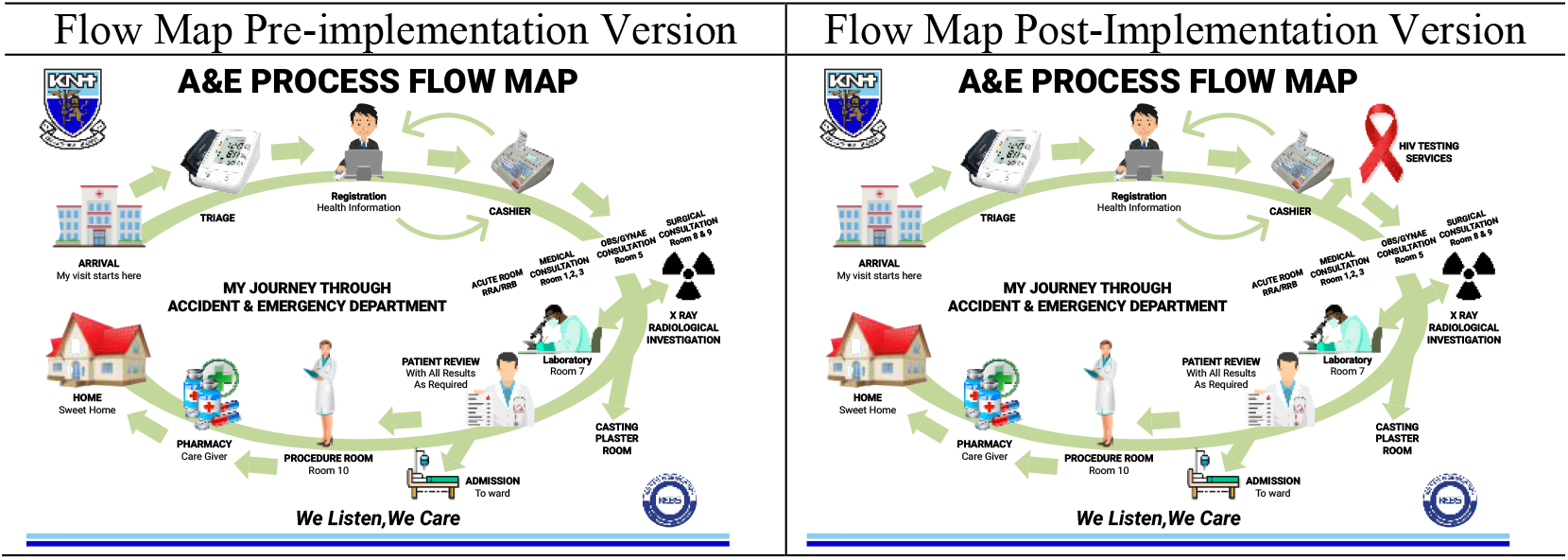

##### HIVST Supply chain

Based on an identified need to ensure continuous access to HIV self-test (HIVST) kits the supply chain was supported during the assessment phase. This was done such that HIVST kits were only provided by study resources in the case that there was an interruption in the standard supply chain. Over the full assessment period an average of 22 HIVST kit per week were supplied to augment access. As increased access for HIVST kit delivery was in alignment with goals from NASCOP the augmentation of this resource within the implementation assessment was deemed to be consistent with the national trajectory for HTS in Kenya.

#### Opportunity

The opportunity domain refers to factors affecting the provision of the behavior ED-HTS delivery based on the physical and social environment that the behaviors must occur within. The primary intervention functions for opportunity to improve HTS were modeling, incentivization and education. Components for the HEATED program for Opportunity were:

##### Promotion of HTS Patient Engagement

To improve the engagement of healthcare workers and patients, and as such the opportunity for HIV services, a single universal screening question was added to the triage process with the goal of ensuring that all patients have an interaction with a care provider on the topic (Triage forms shown below). The question assessed for interest in learning about HTS during the ED care period and encompass a binary (yes or no) response. During the sensitization symposia the healthcare personnel were provide example language on how to ask this however, a standard script was not provided. The patient’s response to this question was be noted in the standard triage documentation which the patient carries throughout their ED course and is reviewed by their clinical care team.

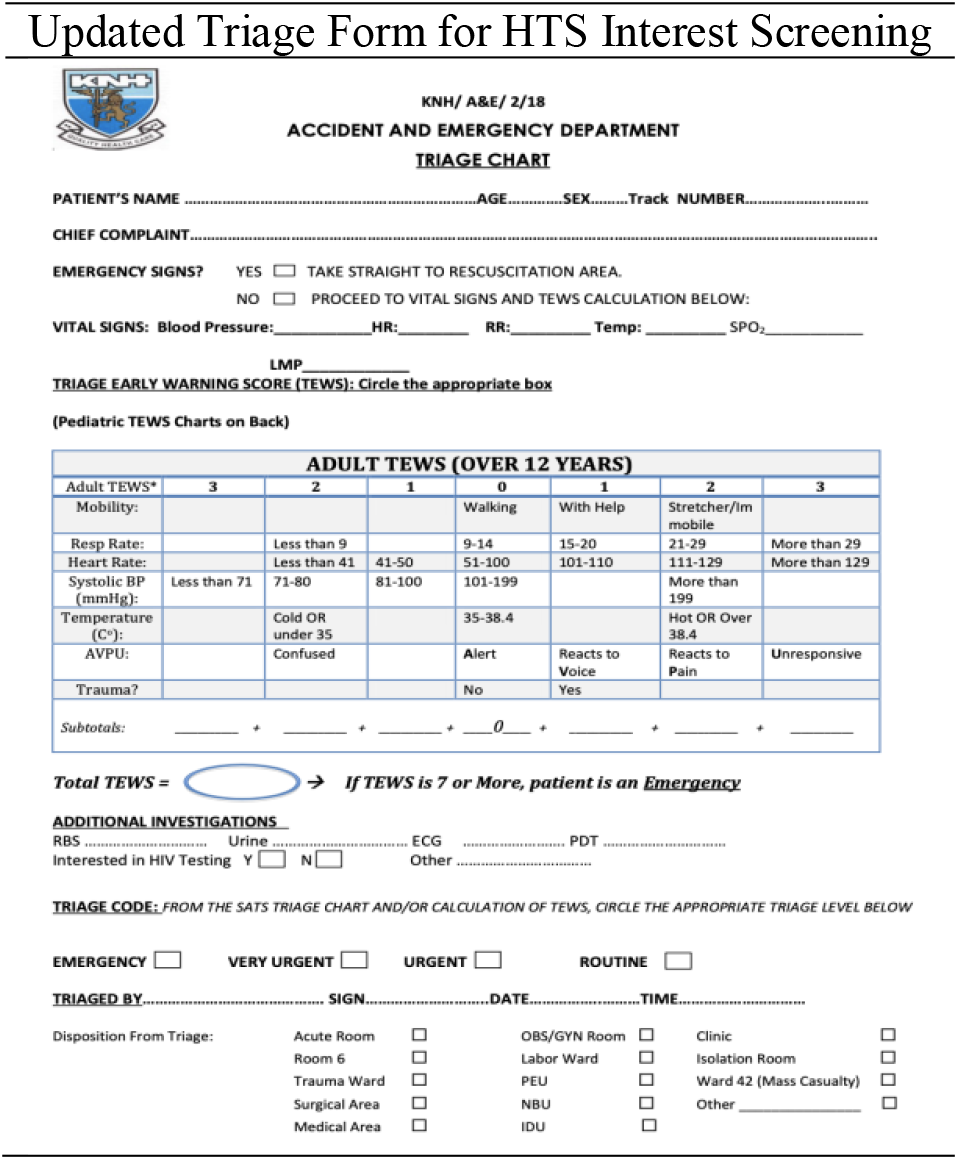

##### Longitudinal Feedback

To improve the social environment and opportunity for HTS the HEAT program provided feedback to normalize and promote HTS at departmental and individual provider levels. This feedback provided updates on HTS metrics including but not limited to: provision of HTS (including HIVST), identification of PLHIV and linkage to care. The feedback was disseminated via digital communication to established ED provider groups on WhatsApp™ every two weeks during the post-implementation period. The feedback was framed in a positive manner and sent by the respective program champions to the nurse, physician and ED-HTS services personnel

#### Motivation

Motivation domain refers to the automatic and reflective processes that affect the stakeholders desire and ability to engage in the health behavior of ED-HTS. The main intervention functions in this domain to improve HTS were healthcare workers drive to deliver HTS (inclusive of conceptual barriers) and patients’ perceptions of the impacts of HTS. Components for the HEATED program for motivation were:

##### Healthcare Worker Champions

To support motivation and engagement in the HEAT program clinical personnel working in the ED were identified who were willing to provide peer-to-peer support within the clinical setting as needed. And through digital communication to ED provider groups on WhatsApp™. This included discussing HTS with ED staff and answering questions that peers may have. For healthcare champions there was one nurse, one physician and the facility director of HTS, these persons were provided a one hour additional training session with the study coordinator to ensure understanding of the HEATED program component and goals.

##### Linkage Peers

A challenge identified for ED-HTS was the difficulty of HIV services personnel to be able to identify patients interested in testing due to overall volume. To support connection of those interested in HTS with the personnel able to provide testing access two linkage peers were placed in the clinical setting for 12 hours a day (7am-7pm). will be placed in the A&E to assist with patient-to-HTS-provider linkage. These personnel were primarily tasked with interacting with patients to review and answer questions about their interest in learning about HTS, and if desired by the patient support their connection to the existing standard ED-HIV services personnel for HTS as appropriate. Although this specific role did not exist in the ED setting prior to the HEATED program there are in place peer-mentors who function primarily to assist with linking patients that test HIV positive to further care. As such the linkage peer role was deemed not to be a substantial alteration in the care delivery process of HTS and was accepted by the KNH HTS program.

##### HIVST Follow up Support

During the qualitative study that there is Substantial attrition of follow up contact for persons who given HIVST kits was identified as a challenge to ED programming. To address this any person receiving HIVST kits were given a NASCOP created and approved informational packet which provides follow up options and mechanisms (see pamphlet below). Additionally, the healthcare worker champion from HTS services provided prompts on follow up goals and department level follow up metrics at standing meetings for the ED-HIV services personnel.

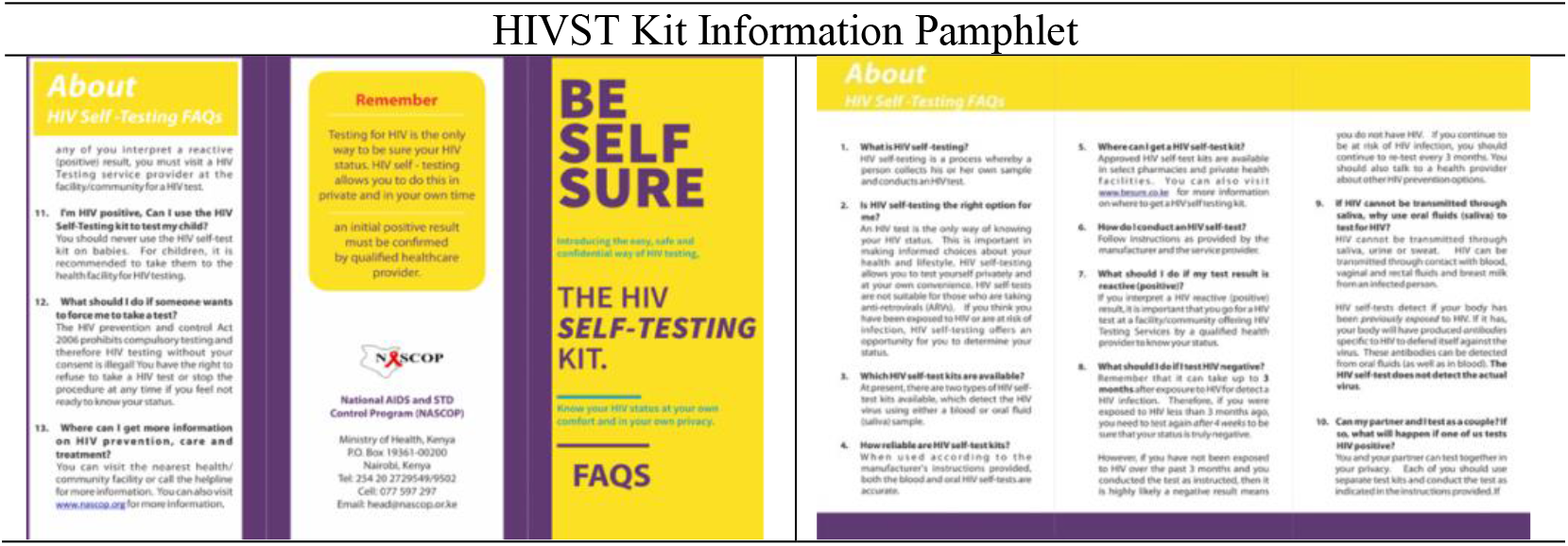

**Supplement 2.**
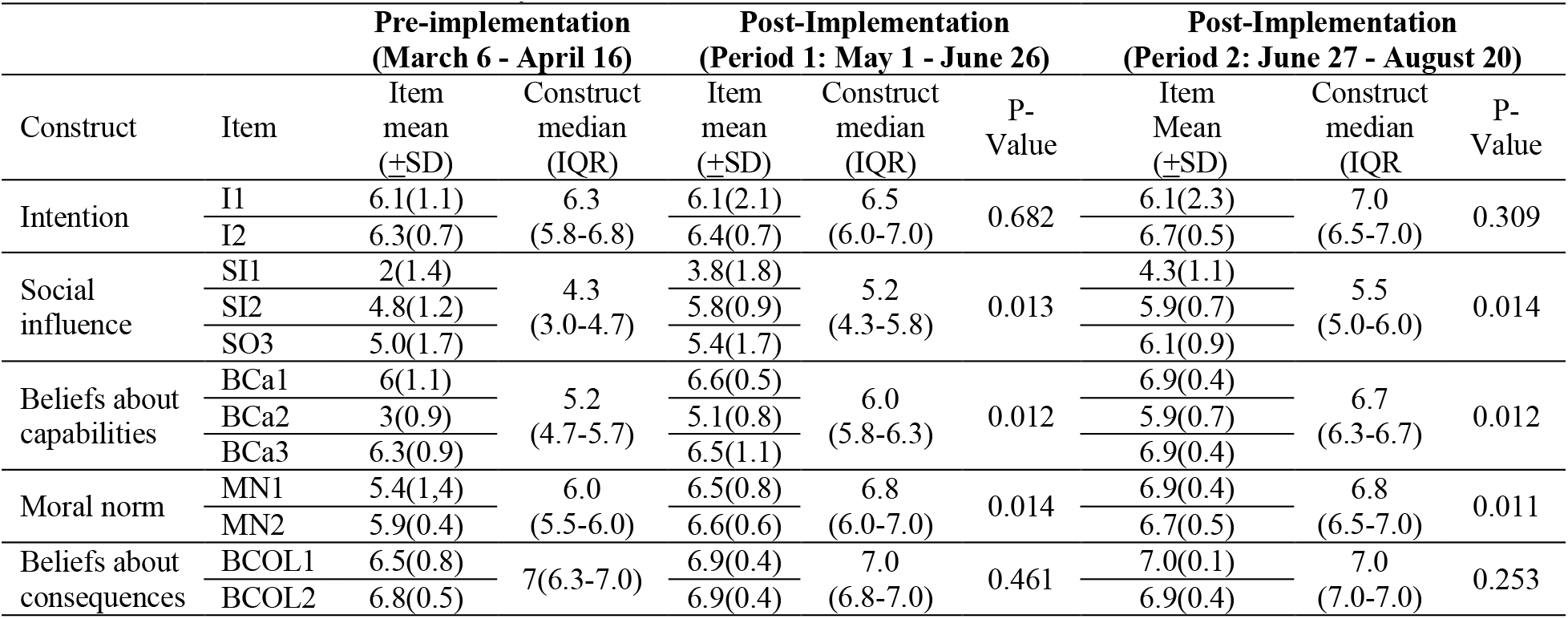
Emergency Department HIV services personnel Continuing Professional Development-Reaction Assessments Across Study Periods.

| Construct | Item | Pre-implementation<br>(March 6 - April 16) |  | Post-Implementation<br>(Period 1: May 1 - June 26) |  | P-Value | Post-Implementation<br>(Period 2: June 27 - August 20) |  | P-Value |
| --- | --- | --- | --- | --- | --- | --- | --- | --- | --- |
| | | Item<br>mean<br>(±SD) | Construct<br>median<br>(IQR) | Item<br>mean<br>(±SD) | Construct<br>median<br>(IQR) | | Item<br>Mean<br>(±SD) | Construct<br>median<br>(IQR) | |
| Intention | I1 | 6.1(1.1) | 6.3 | 6.1(2.1) | 6.5 | 0.682 | 6.1(2.3) | 7.0 | 0.309 |
|  | I2 | 6.3(0.7) | (5.8-6.8) | 6.4(0.7) | (6.0-7.0) |  | 6.7(0.5) | (6.5-7.0) |  |
| Social<br>influence | SI1 | 2(1.4) | 4.3<br>(3.0-4.7) | 3.8(1.8) | 5.2<br>(4.3-5.8) | 0.013 | 4.3(1.1) | 5.5<br>(5.0-6.0) | 0.014 |
|  | SI2 | 4.8(1.2) |  | 5.8(0.9) |  |  | 5.9(0.7) |  |  |
|  | SO3 | 5.0(1.7) |  | 5.4(1.7) |  |  | 6.1(0.9) |  |  |
| Beliefs about<br>capabilities | BCa1 | 6(1.1) | 5.2<br>(4.7-5.7) | 6.6(0.5) | 6.0<br>(5.8-6.3) | 0.012 | 6.9(0.4) | 6.7<br>(6.3-6.7) | 0.012 |
|  | BCa2 | 3(0.9) |  | 5.1(0.8) |  |  | 5.9(0.7) |  |  |
|  | BCa3 | 6.3(0.9) |  | 6.5(1.1) |  |  | 6.9(0.4) |  |  |
| Moral norm | MN1 | 5.4(1.4) | 6.0<br>(5.5-6.0) | 6.5(0.8) | 6.8<br>(6.0-7.0) | 0.014 | 6.9(0.4) | 6.8<br>(6.5-7.0) | 0.011 |
|  | MN2 | 5.9(0.4) |  | 6.6(0.6) |  |  | 6.7(0.5) |  |  |
| Beliefs about<br>consequences | BCOL1 | 6.5(0.8) | 7(6.3-7.0) | 6.9(0.4) | 7.0<br>(6.8-7.0) | 0.461 | 7.0(0.1) | 7.0<br>(7.0-7.0) | 0.253 |
|  | BCOL2 | 6.8(0.5) |  | 6.9(0.4) |  |  | 6.9(0.4) |  |  |

**Supplement 3.** Healthcare Worker Open-ended Feedback on the HIV Enhanced Access Testing in the Emergency Department Program.

| **Vocational Role** | **Feedback Responses (Selected)** |
| --- | --- |
| Nurse | "Identify better resources for (HIV) testing and supporting the manpower helped the emergency department" |
| Nurse | "By sensitizing staff in department on the program of (HIV) testing of the patients the program improved care to those needing it" |
| Nurse | "Testing was easier with the HEAT work, as well as diagnosis and treatment" |
| Nurse | "The program helped get early diagnosis and early treatment hence better outcomes" |
| Physician | "I think that more newly diagnosed HIV patients were captured with the program in the emergency department" |
| Nurse | "HIV testing among trauma patients although improved was not consistent enough" |
| Physician | "The program was implemented but not all the staff were fully sensitized and engaged" |

## Notes

### Competing Interest Statement

The authors have declared no competing interest.

### Funding Statement

This study was funded by National Institute of Allergy and Infectious Diseases (grant number: K23AI145411)

### Author Declarations

The study was approved by the University of Nairobi ethics and research committee (P667/08/2022) and the institutional review board of Rhode Island Hospital (1953237-1). All enrolled participants provided written informed consent.

